# Improving Consistency in Colorectal Cancer Genetic Risk Assessment and Return of Results Through an Integrated Polygenic Risk Score

**DOI:** 10.64898/2026.11.06.26367439

**Authors:** Huy Tran, Ashley J. Mulford, Jun Wei, Zhuqing Shi, Annabelle Ashworth, S. Lilly Zheng, Brian T. Helfand, Peter J. Hulick, Allison DePersia, Grace Suh, Blake Jones, Alan R. Sanders, Jianfeng Xu

## Abstract

**Background:** Published colorectal cancer (CRC) polygenic risk scores (PRS) may demonstrate similar population-level predictive performance but often assign substantially different risk classifications to the same individuals. Such discordance presents challenges for clinical implementation and return of results (RoR).

**Methods:** We developed a stable PRS for CRC (stPRS-CRC) by integrating 28 published PRS using PRSmix in the All of Us (AoU) development cohort (n=321,703). Performance was evaluated in a held-out AoU validation cohort (n=80,423) and independently validated in the Genomic Health Initiative (GHI) (n=17,545). CRC associations were assessed using logistic regression, comparing individuals in the highest PRS decile with the remaining 90%. Individual-level concordance in high-risk classification was evaluated using mean Jaccard indices.

**Results:** Published CRC PRS showed substantial discordance in individual-level high-risk classification despite generally consistent associations with CRC. stPRS-CRC integrated information from multiple published PRS and demonstrated CRC associations comparable to PGS003852, the strongest-performing individual PRS. The odds ratios for stPRS-CRC were 1.94 (95% CI, 1.79–2.11) in AoU development, 2.08 (1.76–2.45) in AoU validation, and 2.08 (1.55–2.79) in GHI. Mean Jaccard indices using stPRS-CRC versus PGS003852 as anchors, each compared with the same 27 published PRS, were 0.29 versus 0.20, 0.43 versus 0.25, and 0.27 versus 0.19, respectively (all *P*<0.001).

**Conclusions:** Integrating multiple published CRC PRS significantly improved individual-level concordance in high-risk classification while maintaining population-level predictive performance. stPRS-CRC may provide a more consistent framework for clinical genetic risk assessment and ROR.

## Introduction

Polygenic risk scores (PRS) have emerged as promising tools for colorectal cancer (CRC) risk assessment, with potential applications to personalized screening and prevention.^1^ By capturing inherited susceptibility associated with common genetic variants, PRS complement rare pathogenic variants (PVs) in hereditary CRC genes, providing a foundation for integrated genetic risk assessment.^2–3^ Our previous studies in the Genomic Health Initiative (GHI), a real-world clinical cohort with genomic and electronic health record (EHR) data, demonstrated the potential utility of inherited genetic risk assessment for identifying individuals at increased CRC risk and informing colonoscopy screening strategies.^4–5^

However, our experience applying two sequentially published CRC PRS (PGS000765 and PGS003852) in the GHI highlighted an important challenge for clinical implementation.^6,7^ Both PRS demonstrated effective population-level risk stratification, with the newer score showing improved predictive performance. Despite these encouraging results, the two PRS assigned discordant risk classifications to some individuals, with a low Jaccard index of 0.21, indicating that only 21% of individuals classified in the highest-risk 10% by either PRS were classified as high risk by both. This distinction between population-level predictive performance and individual-level risk classification consistency is particularly important for return of results (RoR), as replacing an existing PRS with a newer version may alter risk assessments previously communicated to patients and physicians, even though their underlying genetic information remains unchanged.

To address this challenge, we applied PRSmix to develop a stable polygenic risk score (stPRS) framework that integrates information from multiple published PRS.^8,9^ Different PRS may capture overlapping but distinct components of underlying genetic susceptibility, reflecting differences in genetic variants, discovery populations, and development methods.^9–11^ By combining complementary information across existing scores, this approach reduce dependence on any single published PRS and improve consistency in individual-level risk classification while preserving predictive performance. Such consistency may be particularly valuable for clinical implementation and ROR, where maintaining continuity in genetic risk assessment is important.^12–15^

In this study, we developed stPRS-CRC using data from the All of Us (AoU) Research Program and evaluated its predictive performance and individual-level risk classification. We conducted external validation in the real-world GHI cohort. We also systematically examined concordance in high-risk classification across published CRC PRS, with particular attention to implications for personalized CRC screening and sustainable RoR.

## Methods

### Study Design and Overview

This study evaluated whether integrating multiple published CRC PRS could improve consistency in individual-level risk classification while preserving or improving associations with CRC. To minimize potential bias arising from overlap between PRS development and evaluation populations, we selected AoU and GHI, two cohorts that were not major contributors to the GWAS datasets used to develop published CRC PRS. We evaluated 28 published CRC PRS and developed stPRS-CRC in AoU. The AoU cohort was randomly divided into a development set (80%) and a held-out validation set (20%). The development set was used to estimate stPRS-CRC model coefficients, whereas the validation set was reserved for evaluating predictive performance and individual-level risk classification. We also externally validated stPRS-CRC in GHI, a real-world healthcare system biobank with a genetically diverse population.

### Study populations

AoU is a nationwide research program established by the National Institutes of Health that integrates genomic, EHR, and participant-reported data.^16–17^ Participants with available genomic data and CRC phenotype information were eligible for analysis. CRC cases were identified using EHR diagnosis codes corresponding to malignant neoplasms of the colon and rectum (ICD-10 C18–C20), and participants without a documented CRC diagnosis served as controls. The Curated Data Repository (CDR) Version 9 (v9) was analyzed in the study.

The GHI is an ongoing research program and biobank at Endeavor Health that integrates genomic and longitudinal EHR data that contributes to supporting clinical implementation of genetic risk assessment.^4,18^ Participants with available genetic data were included. CRC cases were identified using ICD-10 codes (C18–C20, Z85.038, Z85.04) and ICD-9 codes (153, 154.0, 154.1, V10.05, V10.06 [excluding “anal cancer”]). The last date of HER data access was May 8 2026. GHI served as the primary real-world external validation cohort.

### Published CRC PRS

Candidate CRC PRS were identified from the PGS Catalog.^19^ To support generalizability across genetically diverse populations, we restricted inclusion to PRS developed using multi-ancestry datasets. A total of 28 published CRC PRS met the eligibility criteria and were included in the analysis (**Supplementary Table 1**). None of these PRS was originally developed using AoU or GHI data. Information on the original development populations, genetic variants, weighting methods, and publication dates was obtained from the corresponding publications and PGS Catalog records.

Each PRS was calculated using its published genetic variants and weights.^19^ Variants were harmonized across genomic datasets, and effect allele orientations were verified before score calculation. To account for population structure, raw PRS were adjusted for genetic ancestry principal components (PCs) using the residualization approach described.^20^ The adjusted scores were standardized to have a mean of 0 and standard deviation (SD) of 1 within each analytic population, and percentile ranks were calculated for individual-level comparisons.

### Development of stPRS-CRC

We developed stPRS-CRC by integrating the 28 published CRC PRS using PRSmix.^8.9^ PRSmix uses elastic-net regularization to jointly model multiple PRS, allowing correlated or redundant predictive information to be downweighted while retaining complementary information from individual scores.

Model coefficients were estimated exclusively in the AoU development set using the glmnet R package, with cross-validation for tuning parameter selection. The 28 published PRS were used as candidate predictors to develop a single integrated stPRS-CRC. Once developed, the model coefficients were fixed and applied without modification to the held-out AoU validation set and the independent GHI cohort.

### Population-Level CRC Associations and Predictive Performance

We evaluated the associations with CRC for 28 published PRS and stPRS-CRC across the AoU development and validation sets, as well as in the GHI. The primary population-level analysis compared individuals in the highest 10% of each PRS distribution with the remaining 90%. Associations were estimated using multivariable logistic regression adjusted for age, sex, and ancestry PCs and reported as odds ratios (ORs) with 95% confidence intervals (CIs). The associations of stPRS-CRC with CRC were compared with those of individual published PRS, particularly PGS003852, which demonstrated the strongest association with CRC among the published PRS in the GHI.

As secondary analyses, we assessed associations with CRC using each PRS as a continuous variable (per SD increase) and evaluated predictive discrimination using the area under the receiver operating characteristic curve (AUC), with 95% CIs. To evaluate the incremental predictive value of combining stPRS-CRC and PGS003852, we fitted three multivariable logistic regression models separately in each cohort: Model A included stPRS-CRC, Model B included PGS003852, and Model C included both PRS. All models were adjusted for the same covariates. AUCs were calculated for each model, and pairwise differences were evaluated using DeLong tests.

### Individual-Level Risk Classification

To assess consistency in individual-level risk classification, we evaluated agreement among published CRC PRS and stPRS-CRC. Individuals in the highest 10% of each PRS distribution were classified as high risk. Agreement between high-risk groups was quantified using the Jaccard index, defined as the number of individuals classified as high risk by both scores divided by the number classified as high risk by either score.

We calculated pairwise Jaccard indices among the 28 published CRC PRS. To compare stPRS-CRC with PGS003852, we used the same 27 published comparator PRS, excluding PGS003852. For each anchor score, the mean Jaccard index was calculated across these 27 comparisons. Differences in mean anchored Jaccard indices between stPRS-CRC and PGS003852 were evaluated using participant-level bootstrap resampling, with both mean indices and their difference recalculated in each bootstrap sample to account for dependence among comparisons. These analyses were conducted in the AoU development and validation sets and the GHI.

As secondary analyses, we calculated absolute differences in percentile ranks between pairs of published PRS and examined illustrative plots of individual percentile rankings in the GHI.

### Statistical analysis

Continuous variables were summarized using means and SDs or medians and interquartile ranges (IQR), as appropriate. Categorical variables were summarized using counts and percentages. Associations between PRS and CRC were assessed using multivariable logistic regression. All statistical tests were two-sided, with *P*<0.05 considered statistically significant.

All stPRS-CRC model coefficients were estimated exclusively in the AoU development set and applied without modification to the held-out AoU validation set and the external GHI validation cohort. Statistical analyses were performed using R, including the glmnet package for PRSmix development.

### Ethical Approval

The study was conducted in accordance with applicable institutional review board (IRB) approvals and data-use agreements for AoU. AoU analyses were performed within the program’s authorized research environment. The GHI was approved by the IRB of Endeavor Health. All GHI participants provided informed consent.

## Results

### Study Populations

The study included 419,671 participants across three analytical cohorts, of which 5,148 were CRC cases. AoU included 402,126 participants (4,846 CRC cases), with 21,703 participants (3,899 cases) in the development set and 80,423 participants (947 cases) in the held-out validation set. The GHI included 17,545 participants (302 CRC cases). The cohorts differed in age, sex distribution, genetic ancestry, and CRC prevalence (**Table 1**).

**Table 1.** Characteristics of study cohorts.

|  | **All of Us** |  |  | **GHI** |
|---|---|---|---|---|
|  | Entire cohort | Development | Validation | Entire cohort |
| Subjects, N | 402,126 (100.00) | 321,703 (80.00) | 80,423 (20.00) | 17,545 (100.00) |
| Age at recruitment, median (IQR) | 46 (31-58) | 46 (31-58) | 46 (31-58) | 57 (43-67) |
| Sex, male, N (%) | 156,605 (38.94) | 125,425 (38.99) | 31,180 (38.77) | 6,208 (35.38) |
| European-like genetic ancestry*, N (%) | 226,564 (56.34) | 181,239 (56.34) | 45,325 (56.36) | 12,542 (71.48) |
| CRC cases, N (%) | 4,846 (1.21) | 3,899 (1.21) | 947 (1.18) | 302 (1.72) |
| Age at diagnosis, median (IQR) | 61 (53-70) | 61 (52-70) | 62 (53-70) | 65 (57-72) |

\*European-like genetic ancestry was defined as a combined assignment probability ≥0.75 for the non-Finnish European (NFE), Finnish (FIN), and Ashkenazi Jewish (ASJ) reference groups

### Associations and Concordance of Published CRC PRS in Real-World GHI

To assess population-level CRC associations and individual-level concordance among published PRS in a real-world setting, we evaluated 28 published CRC PRS in the GHI. Comparing individuals in the highest 10% of each PRS with the remaining 90%, 27 of the 28 PRS had ORs greater than 1, and 16 were significantly associated with CRC (*P*< 0.05) (**Figure 1a**). PGS003852 showed the strongest association, with an OR of 2.17.

**Figure 1.**
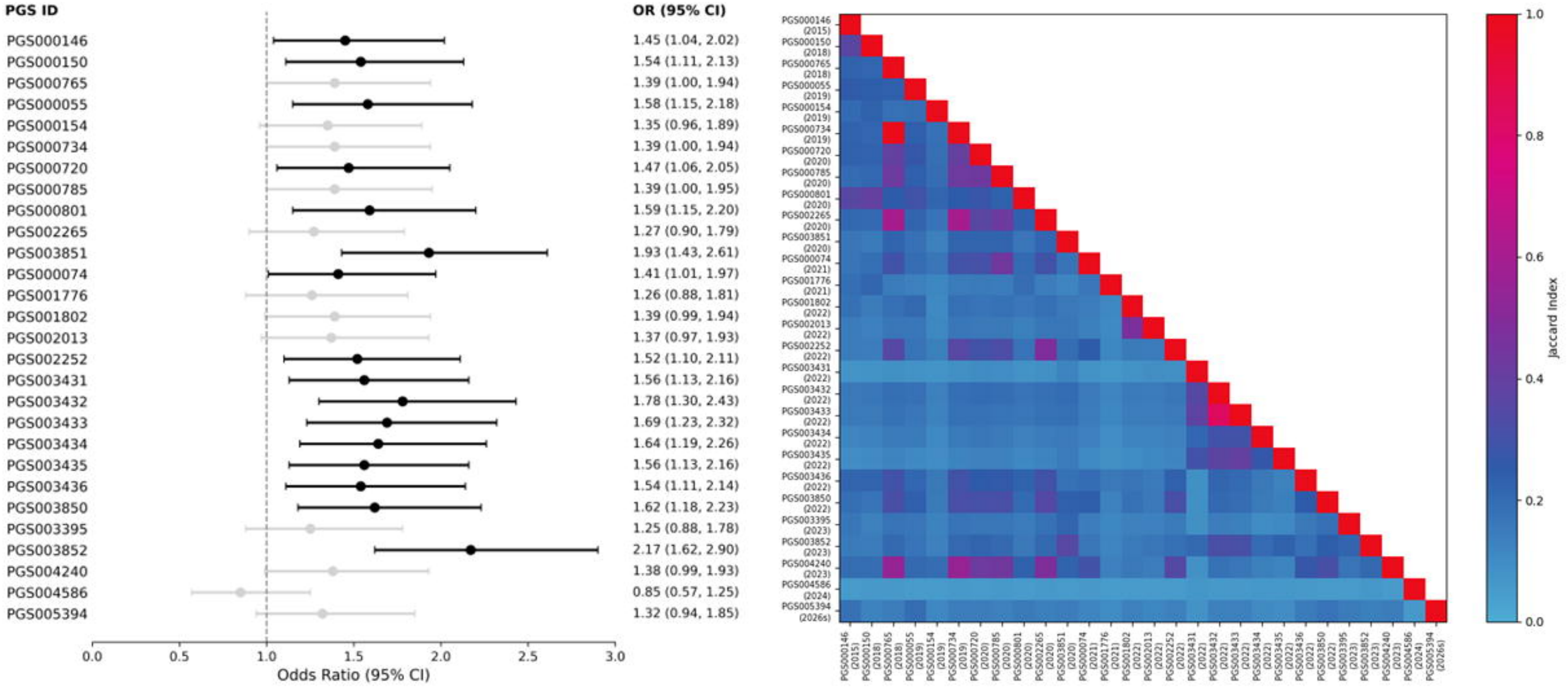
Predictive performance and individual-level risk classification concordance of 28 published CRC PRS in GHI. (a) Forest plot showing odds ratios (ORs) and 95% confidence intervals (CIs) for CRC among individuals in the highest 10% of each PRS compared with the remaining 90%. (b) Heatmap showing pairwise Jaccard indices among the 28 published CRC PRS based on classification of individuals in the highest 10% of genetic risk. The Jaccard index represents the proportion of individuals classified as high risk by both PRS among those classified as high risk by either PRS. Higher values indicate greater concordance in individual-level risk classification.

Although many published PRS were associated with CRC at the population level, they showed substantial variation in individual-level high-risk classification. Across 378 pairwise comparisons, Jaccard indices for the highest 10% risk group ranged from 0.05 to 0.83, with a median of 0.16 (IQR, 0.13–0.22) (**Figure 1b**).

### Development of stPRS-CRC in AoU

We developed stPRS-CRC in the AoU development cohort by integrating 28 published CRC PRS using PRSmix. The resulting model assigned nonzero coefficients to 12 PRS, with PGS003852 receiving the largest coefficient (0.12), substantially higher than those of the other PRS (**Supplementary Table 2**).

At the population level, stPRS-CRC was significantly associated with CRC, with an OR of 1.94 (95% CI, 1.79–2.11) for individuals in the highest 10% compared with the remaining 90% (**Table 2**). The association of stPRS-CRC with CRC was similar in magnitude to that of PGS003852; OR of 1.96 (95% CI, 1.81–2.13), and stronger than that of most individual PRS (**Supplementary Table 3**). In secondary analyses treating PRS as continuous variables, stPRS-CRC was also associated with CRC, with an OR of 1.38 (95% CI, 1.34–1.43) per SD increase, compared with 1.40 (95% CI, 1.36–1.45) for PGS003852 (**Supplementary Table 4**).

**Table 2.** Population-level predictive performance and individual level concordance.

|  | **OR (95% CI) [Categorical]** |  | **Mean Jaccard index anchored by** |  |  |  |
|---|---|---|---|---|---|---|
| **Cohort** | stPRS | PGS003852 | stPRS | PGS003852 | Δ (95% CI) | P-value |
| Development cohort |  |  |  |  |  |  |
| AoU-development cohort | 1.94 (1.79, 2.11) | 1.96 (1.81, 2.13) | 0.29 | 0.20 | 0.09 (0.088, 0.091) | <0.001 |
| Validation cohorts |  |  |  |  |  |  |
| AoU-validation cohort | 2.08 (1.76, 2.45) | 2.03 (1.72, 2.40) | 0.43 | 0.25 | 0.18 (0.172, 0.179) | <0.001 |
| GHI | 2.08 (1.55, 2.79) | 2.17 (1.62, 2.90) | 0.27 | 0.19 | 0.08 (0.073, 0.084) | <0.001 |

Note: Adjusted for age, sex, and PCs.

At the individual level, stPRS-CRC demonstrated greater concordance in high-risk classification across published PRS. Using the same 27 comparator PRS, the mean anchored Jaccard index was 0.29 for stPRS-CRC, compared with 0.20 for PGS003852, *P*<0.001 (**Table 2**).

### Validation of stPRS-CRC in AoU and GHI

We evaluated stPRS-CRC in the held-out AoU validation cohort and the independent GHI cohort. At the population level, stPRS-CRC was significantly associated with CRC in both validation cohorts, with ORs of 2.08 (95% CI, 1.76–2.45) and 2.08 (95% CI, 1.55–2.79) for individuals in the highest 10% compared with the remaining 90% in the AoU validation cohort and the GHI, respectively (**Table 2**). Its associations with CRC were similar in magnitude to those of PGS003852 across the two cohorts and stronger than those of most other individual PRS (**Supplementary Table 5**). In secondary analyses treating PRS as continuous variables, ORs were similar between stPRS-CRC and PGS003852 across the two validation cohorts (**Supplementary Table 4**).

At the individual level, stPRS-CRC demonstrated greater concordance with published PRS in high-risk classification across both validation cohorts. Using the same 27 comparator PRS, the mean anchored Jaccard indices for stPRS-CRC were 0.43 and 0.27 in the AoU validation cohort and the GHI, respectively, compared with 0.25 and 0.19 for PGS003852, both *P*<0.001 (**Table 2**).

Further comparison of stPRS-CRC and PGS003852 suggested that the two scores capture complementary information about CRC risk. Across all three analytical cohorts, individuals classified as lower risk by both PRS had the lowest observed CRC prevalence, whereas those classified as high risk by both had the highest prevalence (2.21–2.46 times higher). Individuals classified as high risk by only one PRS had intermediate prevalence (1.35–2.14 times higher), with generally similar estimates regardless of which PRS identified them (**Supplementary Table 6**).

In secondary analyses, we evaluated whether combining stPRS-CRC and PGS003852 improved predictive discrimination compared with either PRS alone. The combined model showed modest improvements in AUC, although the magnitude and statistical significance varied across cohorts (**Supplementary Table 7**).

### Additional Analyses of Individual-Level Risk Classification

To assess variation in risk classification across the entire spectrum of participants, rather than only those classified as high risk, we compared individual percentile rankings across the 28 published CRC PRS. In the GHI, median absolute differences in risk percentile ranks across PRS pairs ranged from 2 to 30 percentile points, with an overall median of 21 (**Supplementary Figure 1a**). Illustrative plots of percentile rankings for 10 randomly selected individuals further demonstrated variability in risk assignment across published PRS (**Supplementary Figure 1b**).

## Discussion

In this study, we developed stPRS-CRC by integrating 28 published CRC PRS and evaluated its performance in three analytical cohorts from AoU and the GHI. Three principal findings emerged. First, although many published CRC PRS were significantly associated with CRC, they showed substantial disagreement in individual-level risk classification. Second, stPRS-CRC demonstrated significantly higher mean concordance with published PRS than PGS003852, the strongest-performing individual PRS, in all three cohorts. Third, stPRS-CRC maintained CRC risk associations comparable to those of PGS003852. Together, these findings demonstrate that integrating multiple published PRS can improve individual-level concordance while maintaining population-level predictive performance.

The substantial individual-level discordance among published CRC PRS, despite their generally consistent population-level associations with CRC, is not unexpected but has important practical implications.^7,21,22^ Similar inconsistencies in individual-level risk classification have been reported for PRS in other diseases, but to our knowledge have not been systematically characterized for CRC.^9,23,24^ One possible explanation is that different PRS capture partially overlapping but distinct components of inherited genetic susceptibility.^9,25^ Differences in the genetic variants included,^23^ their estimated effect sizes,^26^ the ancestry composition of development cohorts,^7,27^ and the methods used for PRS construction may all contribute to this discordance.^22,28^ Consequently, PRS with comparable population-level predictive performance may assign substantially different risk estimates to the same individual.^9,23,24^ This distinction is particularly important for clinical implementation, where risk assessment and subsequent recommendations are made at the individual rather than population level.^15,21,23,26,29^ Such disagreement would create uncertainty for physicians and patients, particularly when newer PRS replace existing ones.^9,25,30^

Our findings suggest that integrating multiple PRS offers a potential approach to addressing this challenge. Rather than selecting a single published PRS based solely on predictive performance, stPRS-CRC combines information across scores using PRSmix.^8,9^ Notably, PGS003852 received the largest coefficient, indicating that stPRS-CRC retained substantial information from the strongest individual PRS while incorporating complementary information from others. The consistently higher mean Jaccard index for stPRS-CRC across all three analytical cohorts supports the potential of this approach to improve individual-level concordance and reduce dependence on any single published score. Furthermore, individuals classified as high risk by both stPRS-CRC and PGS003852 had the highest observed CRC prevalence, whereas those identified by only one score had similarly elevated intermediate prevalence. These findings suggest that the two PRS capture overlapping but partly complementary information about inherited CRC susceptibility.

These findings have important implications for clinical implementation and RoR. By improving consistency in individual-level risk classification, stPRS-CRC should provide a more durable framework for genetic risk assessment as new PRS become available.^9^ Integrating complementary genetic information should also support more comprehensive risk assessment and personalized CRC screening strategies.^21,22,29^ However, combining stPRS-CRC and PGS003852 yielded only modest and inconsistent improvements in AUC compared with PGS003852 alone (**Supplementary Table 7**), suggesting that the primary potential advantage of stPRS-CRC lies in improving consistency rather than predictive discrimination. Whether these benefits translate into improved clinical decision-making and screening outcomes requires further evaluation.

An additional potential advantage of this approach is its adaptability over time. As new CRC PRS are developed and validated, they could be incorporated into updated versions of stPRS-CRC rather than replacing the existing score with each newly published PRS.^9^ This approach would facilitate continuity in genetic risk assessment across successive generations of PRS.^9,13^ Although the potential to reduce reclassification during score updates can be evaluated retrospectively, prospective studies will be important to understand the clinical implications of reclassification for physicians and patients.^31^

Several limitations should be acknowledged. First, our assessment of stability was based on concordance across existing PRS rather than longitudinal changes in risk classification following score updates. Second, higher concordance does not necessarily indicate greater predictive accuracy or clinical utility.^9,23,27^ Third, additional studies are needed to evaluate performance across diverse ancestry groups and to determine how improved consistency influences clinical decision-making and patient understanding.^7,11,14^

In conclusion, stPRS-CRC achieved greater individual-level concordance with published CRC PRS while maintaining predictive performance comparable to the strongest individual score. This approach should provide a more consistent and adaptable framework for integrating polygenic risk assessment into clinical practice.

## Data Availability

AoU data are available to approved researchers through the All of Us Researcher Workbench. This study used the Controlled Tier Dataset, version 9. For GHI, the data supporting the findings of this study are available from Endeavor Health but restrictions apply to the availability of these data due to institutional and participant privacy considerations. De-identified data may be made available from the corresponding author upon reasonable request and with appropriate institutional approvals.

---

**Supplementary Figure 1. Individual-level variation in risk classification across 28 published colorectal cancer polygenic risk scores in the Genomic Health Initiative (GHI) cohort**. (a) Heatmap showing pairwise median absolute differences in percentile ranks among the 28 published colorectal cancer polygenic risk scores (PRS). (b) Variation in percentile ranks across the 28 PRS for 10 randomly selected individuals, illustrating differences in individual-level risk classification.

## Author Contributions

*Concept and design*: Xu J and Tran H

*Data analysis*: Tran H, Shi Z, Mulford AJ, Wei J, Sanders AR

*Manuscript draft*: Xu J and Tran H

*Critical revision of the manuscript for important intellectual content*: All co-authors

*Supervision*: Xu J and Sanders AR

## Acknowledgements

We thank the study subjects and researchers of the National Institutes of Health’s AoU Research Program, as well as the participants of the GHI. We are grateful to the Ellrodt-Schweighauser family for establishing Endowed Chair of Cancer Genomic Research (Xu) and Davis family for the Davis Family Center for Preventive Genomics.

