## Supplementary figures and images for "Improving Consistency in Colorectal Cancer Genetic Risk Assessment and Return of Results Through an Integrated Polygenic Risk Score"

### Supplemental Figure

#### Slide 1
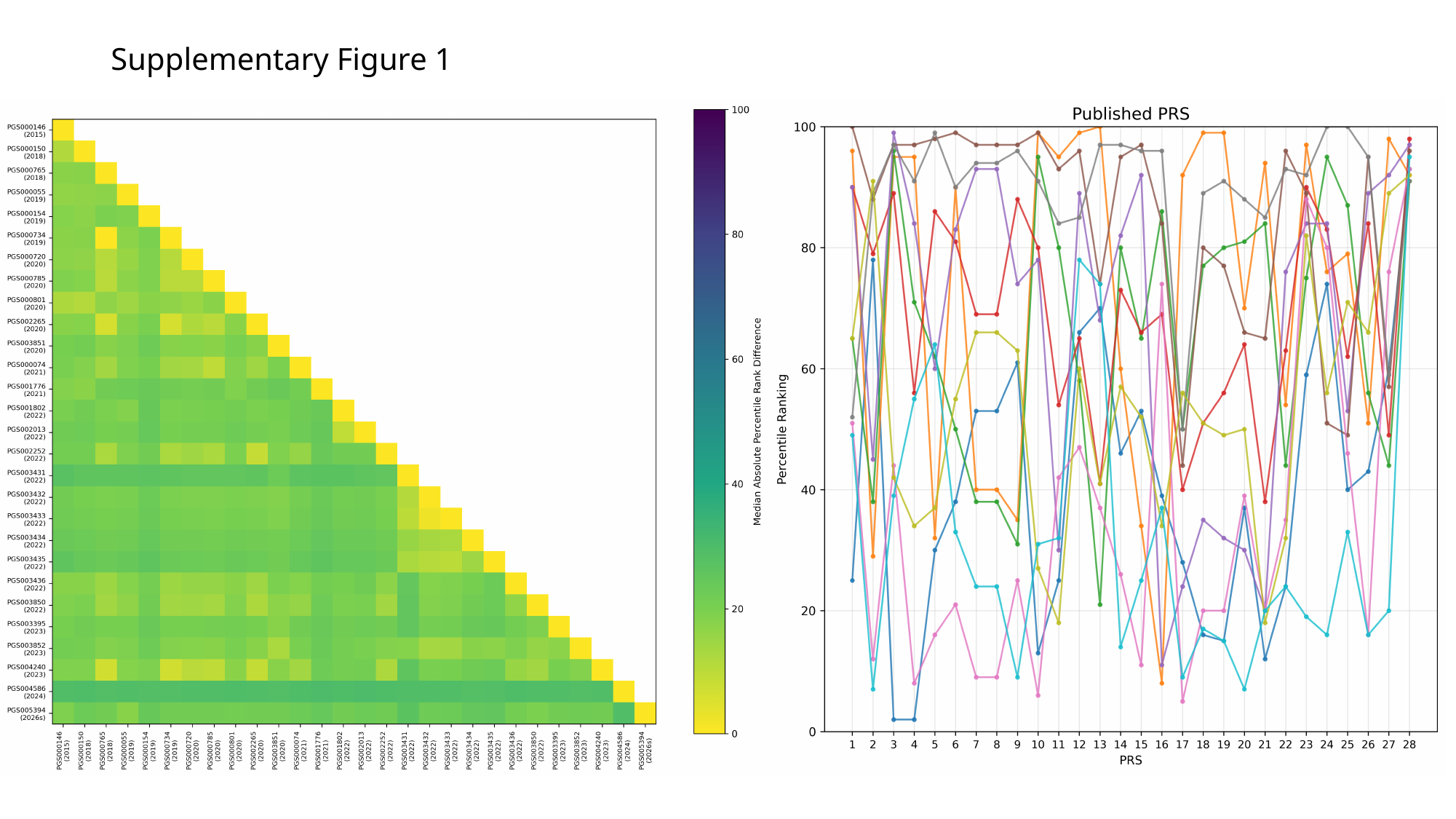

##### Supplementary Figure 1
